# Filtration Performance Degradation of In-Use Masks by Vapors from Alcohol-Based Hand Sanitizers and the Mitigation Solutions

**DOI:** 10.1101/2020.11.01.20223982

**Authors:** Weidong He, Yinghe Guo, Jingxian Liu, Yang Yue, Jing Wang

## Abstract

How often does one perform hand disinfection while wearing a mask? In the current COVID-19 pandemic, wearing masks and hand disinfection are widely adopted hygiene practices. However, our study indicated that exposure to the vapors from alcohol-based sanitizers during hand disinfection might degrade the filtration performance of the in-use masks, and the degradation worsened with the increasing number of hand disinfection. After five times of hand disinfection, the filtration efficiencies of surgical masks decreased by >8% for 400 and 500nm particles and by 3.68±1.83 % for 1μm particles. This was attributed to the dissipation of electrostatic charges on the masks when exposed to the alcohol vapor generated during hand disinfection. Simple practice of vapor-avoiding hand disinfection could mitigate the effects of alcohol vapor, which was demonstrated on two brands of surgical masks. The vapor-avoiding hand disinfection is recommended to be included in the hygiene guide to maintain the mask performance.

**Graphic abstract:** 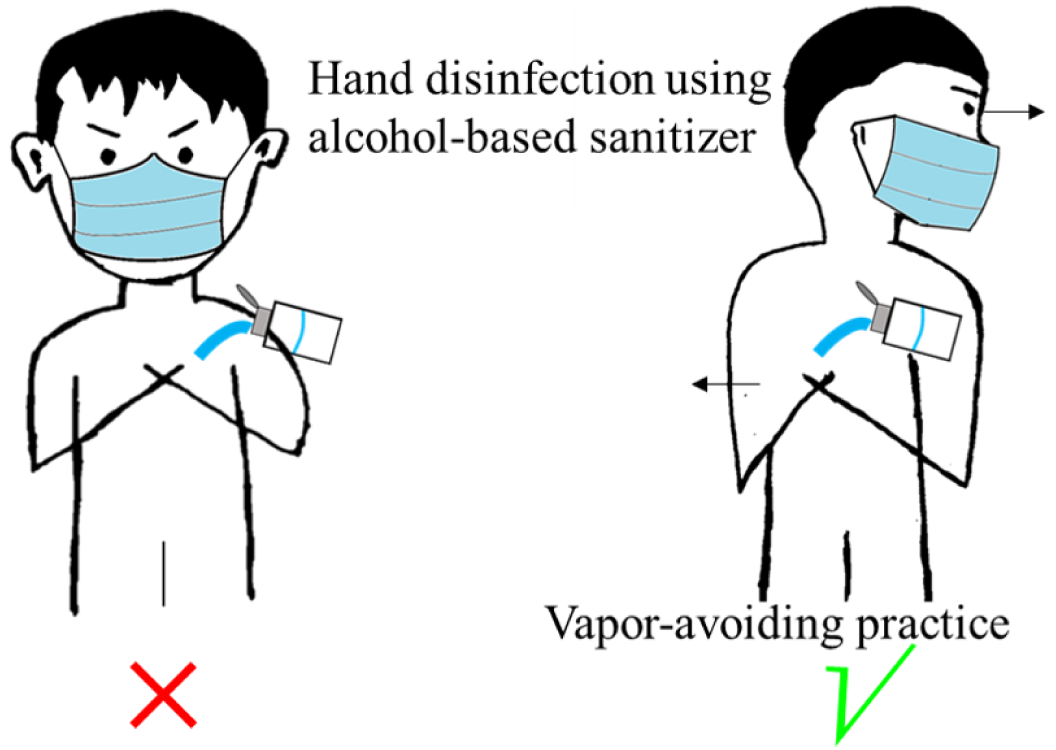

---

The COVID-19 pandemic is raging and many countries are suffering the second wave. Compared with the early stage of the pandemic outbreak, more comprehensive approaches, including contact tracing, quarantine, physical distancing, hand hygiene, and masks, have been proposed to slow down the spread of COVID-19.^1^ Nevertheless, the number of new infections per day has constantly increased since October 2020.^2^ In order to contain the second wave of the pandemic and keep businesses open, regular hand disinfection and mandatory face masks in public places have been ordered or recommended in the latest anti-COVID measures by most countries.^3-5^

A number of studies showed that wearing masks in public could prevent interhuman transmission effectively.^6,7^ In order to solve the mask shortage, various works related to mask regeneration and alternative materials for masks were carried out.^8,9^ Simultaneously, it is confirmed that alcohol-based sanitizers can inactivate the SARS-CoV-2 virus, and they are recommended by the World Health Organization (WHO).^10,11^ Regular hand disinfection and wearing masks in public places will be a necessary part of our life in the foreseeable future. However, using the alcohol-based sanitizers for hand disinfection may degrade the filtration performance of masks. Organic solvents including alcohol-based agents degraded the filtration efficiencies of electrostatic filters by dissipating the electrostatic charges.^12,13^ Ethanol was not recommended for mask regeneration, although it was capable of efficient microbial inactivation.^8^ Many hand sanitizers on the market are alcohol-based. Although the alcohol-based sanitizers would not directly contact the masks worn by the users during the hand disinfection, the vapors of alcohol-based sanitizers could dissipate the charges on the masks, finally leading to insufficient protection for the mask wearers.

In the present work, the effects of hand disinfection using alcohol-based sanitizers on filtration performances of cotton masks, surgical masks, and N95 respirators worn by the users were investigated. The selected masks were shown in Figure S1. Both the electrostatic potentials and filtration efficiencies for 50 nm to 3 μm particles of the masks were measured to evaluate the filtration performance of masks by the setups showed in Figure S2 and Figure S3. The particle sizes were selected according to the report that SARS-CoV-2 aerosols were found in the size range of 250-1000 nm,^14^ and the test standard (EN 14683) for surgical masks is at 3 μm particles. The size distributions of monodisperse particles used for the filtration test could be found in Figure S4. The dependence of the degradation effect on the number of performed hand disinfection was investigated. In addition, we proposed a simple practice for vapor-avoiding hand disinfection to mitigate the effects of alcohol-based sanitizers on mask filtration performance.

WHO published a guide for the detailed hand disinfection steps, but the position of hands during hand rubbing was not mentioned.^15^ Herein, the WHO recommended hand disinfection steps were employed. The duration of the entire procedure was 20-30 seconds. Meanwhile two types of practices featuring different hand and face positions were studied. In one type of practice illustrated on the left of Figure 1, the volunteers placed hands between the abdomen and chest, which was named as the common hand disinfection in the present study. In the other type of practice, to avoid inhaling the sanitizer vapor, the volunteers placed hands on one side of the body and turned the head to the opposite side, as shown on the right in Figure 1. The second practice was named as the vapor-avoiding hand disinfection. For comparison, the performances of brand new masks and masks worn by the volunteers for 5 hours were tested. Common hand disinfection and vapor-avoiding hand disinfection using 60 – 80% 2-Propanol were performed up to 10 times, then the part near the nose/mouth area of the mask where the inhaled airflow was most concentrated was cut out for the performance test. In the figures, “No HD” indicates the mask was worn 5 hours without hand disinfection; “HD x n” indicates that n times of hand disinfection were performed during the 5 hours when the mask was worn.

**Fig. 1.**
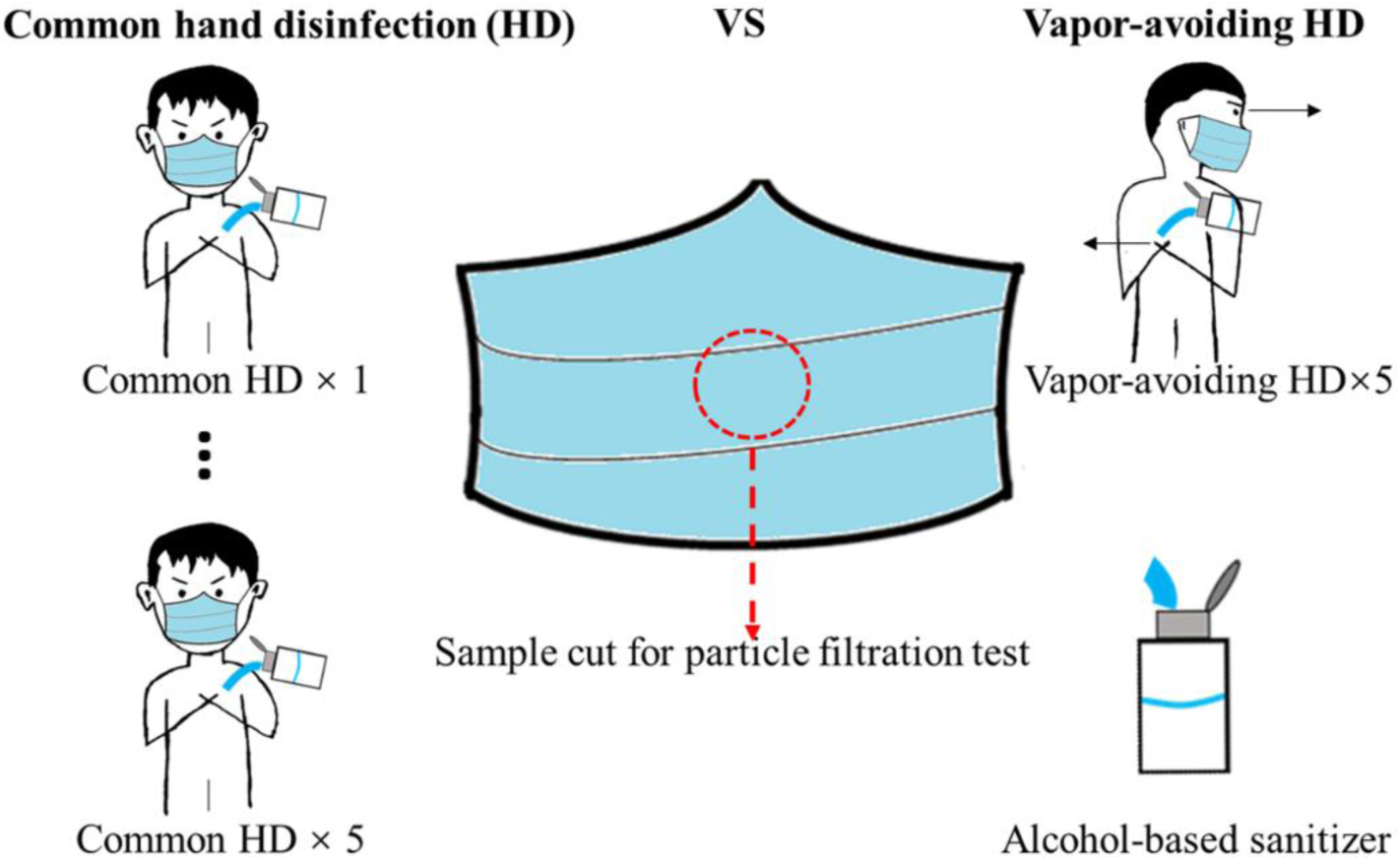
Using alcohol-based sanitizer for common and vapor-avoiding hand disinfection.

A type of cotton mask on the Swiss market was selected to be evaluated (photo in Figure S1). Because the filtration efficiencies of the selected cotton mask for 50-800 nm particles were very low (about 10-20%, Figure S5), we only used the total filtration efficiency of the cotton mask for the polydisperse NaCl particles (Figure S6) to evaluate the effect of exposure to sanitizer vapor. The cotton mask consisted of textile fabric, and its particle capture function only depended on the physical structure instead of electrostatic property. Both the filtration efficiencies and electrostatic potential of the cotton mask had no change after 5 times of common hand disinfection (Figure 2 and Figure S7).

**Fig. 2.**
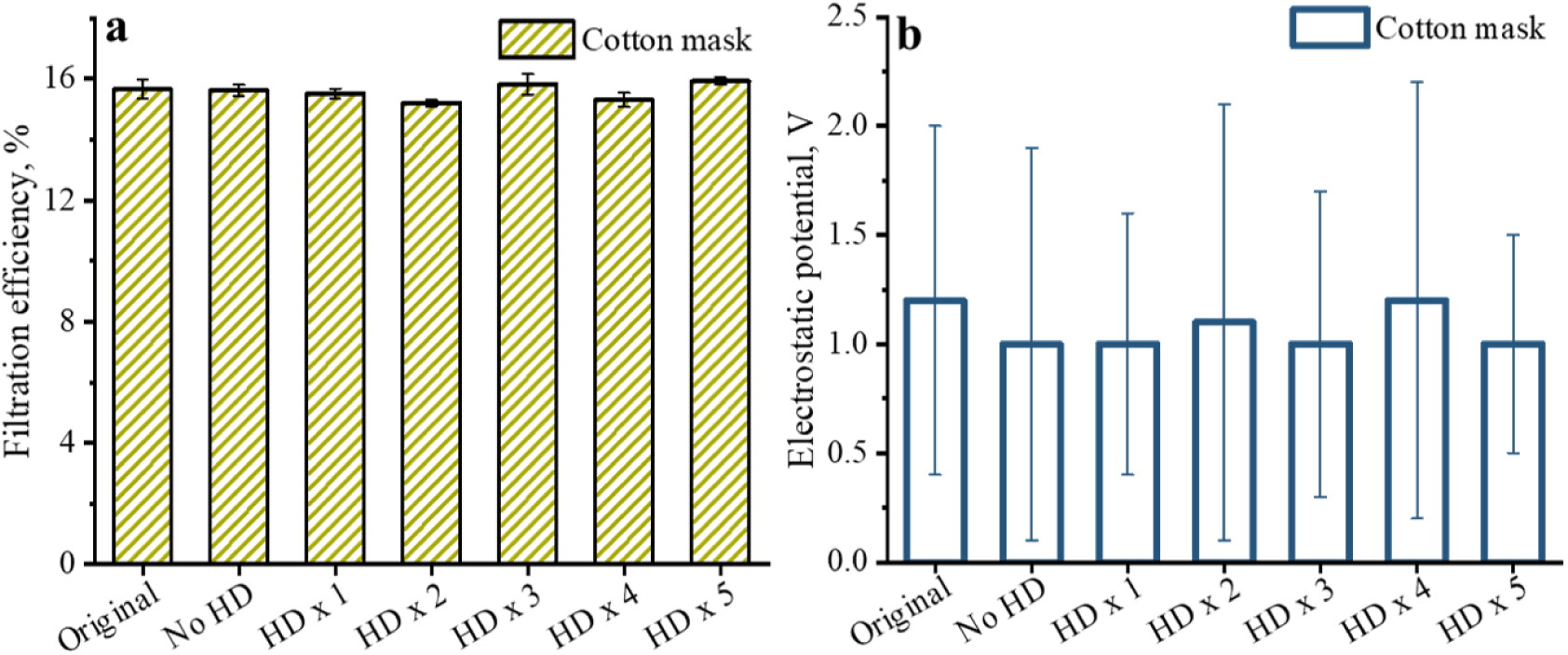
Filtration efficiencies and electrostatic potentials of brand new and used cotton masks without and with several times of common hand disinfection (HD); a) Filtration efficiency; b) electrostatic potential.

For a type of selected surgical masks (brand 1), the average electrostatic potential decreased as the number of common hand disinfection increased (Figure 3a). A statistically significant degradation of the electrostatic potential occurred when the number of hand disinfection increased to 4 times or more (Table S1). The electrostatic potential of all tested N95 respirators had no statistically significant difference after common hand disinfection up to 10 times, which indicated that using alcohol-based sanitizers would not influence the electrostatic property of N95 respirators under the experimental conditions of the present study (Figure 3e).

**Fig. 3.**
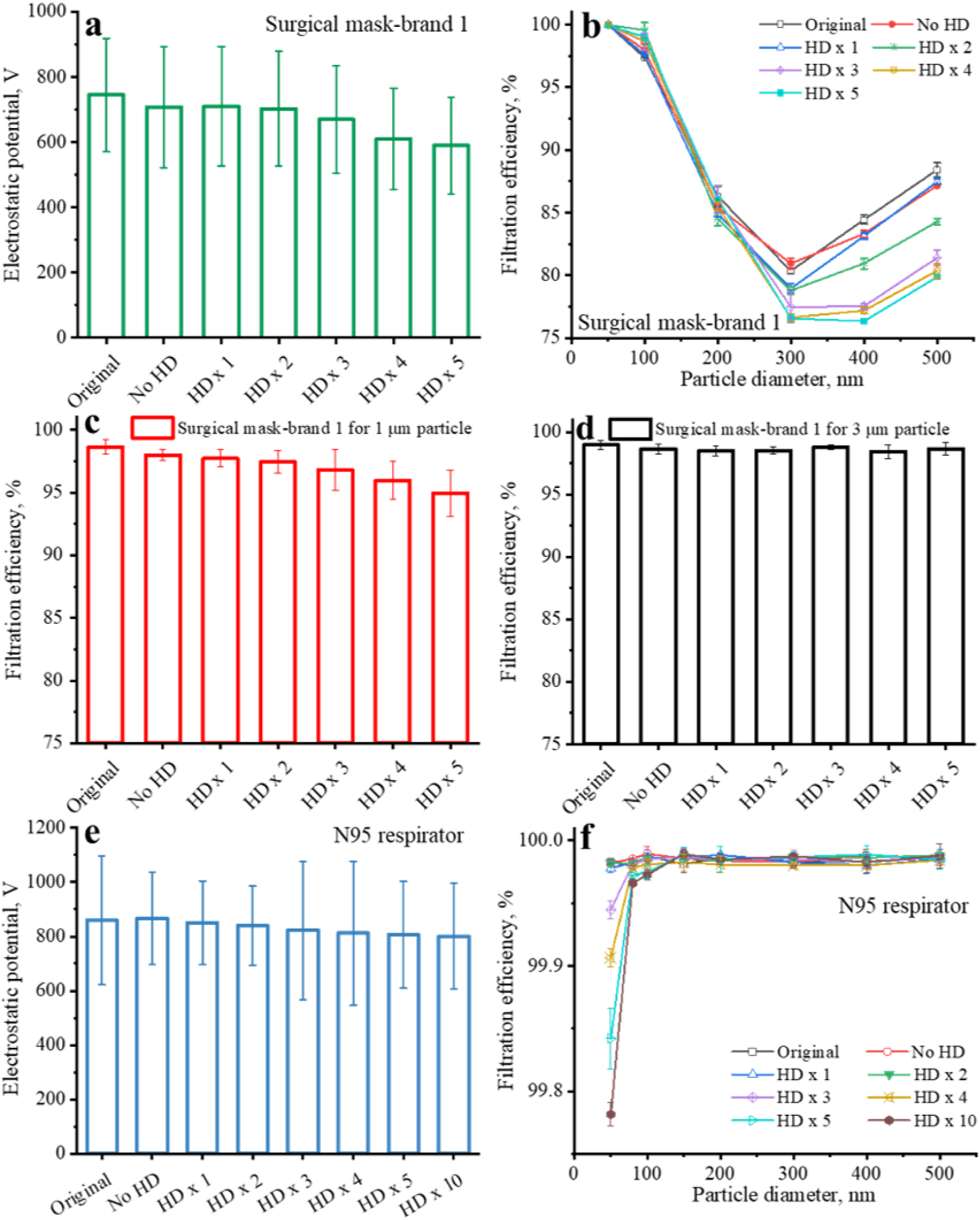
Electrostatic potentials and filtration efficiencies of brand new and used surgical masks (brand 1) and N95 respirators without and with several times of common hand disinfection (HD): a), b), c), and d) surgical mask; e) and f) N95 respirator.

As shown in Figure 3b, the filtration efficiencies of the surgical masks (brand 1) had almost no change after 5 h usage without hand disinfection. In comparison, 1.41±0.41% degradation of the filtration efficiency for 300 nm particles was observed for the surgical masks with 1 time of common hand disinfection. After 2 times of common hand disinfection, the filtration efficiencies of the surgical masks for 400 and 500 nm particles decreased from 84.43±0.36% and 88.41±0.56% to 80.95±0.43% and 84.28±0.24%, respectively. Consistent with the drop in the electrostatic potential, the filtration efficiencies of the surgical masks decreased as the number of common hand disinfection increased (Figure 3b). After 5 times of common hand disinfection, the degradation of filtration efficiency for 300 nm particles exceeded 3%, and more than 8% filtration efficiency degradation for 400 and 500 nm particles was observed. After 4 and 5 times of common hand disinfection, the filtration efficiency for 1 μm particles decreased from 98.61±0.57% to 95.96±1.49% and 94.93±1.83%, respectively (Fig. 3c). The filtration efficiency for 3 μm particles was not affected (Fig. 3d). The different alcohol vapor effects for various particle sizes were attributed to the underlying filtration mechanisms: electrostatic capture plays a significant role for small particles in the sub-micron range, whereas interception and inertial impaction dominate for particles above several micrometers.^16^ SARS-CoV-2 aerosols were found in the size range of 250-1000 nm.^14^ Therefore, the filtration efficiency degradation of surgical masks after common hand disinfection for several times would diminish the protection for the mask wearers who are exposed to airborne SARS-CoV-2 aerosols. Common hand disinfection up to 10 times had no obvious influence on the filtration efficiencies of the N95 respirator for particles in the range of 80-500 nm (Figure 3f). The filtration efficiency of the N95 respirator for 50 nm particles dropped with the increasing number of common hand disinfection. After 10 times of common hand disinfection, the filtration efficiency of the N95 respirator for 50 nm particles decreased slightly from 99.98±0.003% to 99.78±0.01%, which could still provide high level protection.

The charge de-trapping of electrostatic filters induced by alcohol vapor was the main reason of the filtration efficiency degradation, which was shown in our previous study.^12^ Actually, alcohol vapor treatment is a standard method in ISO/DIS 16890-1 to discharge electret filters and has been widely used in previous studies.^17,18^ It has been noticed that the same common hand disinfection exhibited different influences on surgical masks and N95 respirators, which might be attributed to the different structures of these two types of masks (Figure S8). The particle capture function of both the surgical mask and N95 respirator depends on the inner layer which usually consists of charged PP (polypropylene) melt-blown nonwoven. First, the outermost layer of the N95 respirator was thicker than that of the surgical mask. It is more difficult for the alcohol vapor to penetrate into the inner layer of the N95 respirator. Second, the surgical mask had a single charged PP melt-blown nonwoven inner layer, whereas the N95 respirator possessed multiple nonwoven layers. Although the original electrostatic potentials of two types of masks were similar, the charge amount throughout the entire N95 respirator was higher than the surgical mask. In other words, the alcohol vapor dose from hand disinfection in the present study only dissipated a small percentage of the charges on the N95 respirator, and was not enough to induce notable degradation of the electrostatic potential or filtration efficiency. The vapors from alcohol-based sanitizers during hand disinfection presented more influence on the surgical mask than the N95 respirator. Since surgical masks are widely used by general public now, appropriate mitigation strategies are therefore needed.

Herein, we proposed a vapor-avoiding strategy to mitigate the mask efficiency degradation induced by hand disinfection using alcohol-based sanitizers. The surgical mask-brand 1 used in the above experiments, and another commercial surgical mask-brand 2 were tested to evaluate the vapor-avoiding hand disinfection method. For surgical mask-brand 1, only ∼1% degradation of filtration efficiency for 400 nm particles was observed after 5 times of vapor-avoiding hand disinfection (Figure 4a). The filtration efficiency for 1 μm particles was also maintained when applying vapor-avoiding hand disinfection (Figure 4c). In comparison, the degradation of filtration efficiencies for both 400 and 500 nm particles exceeded 8% after 5 times of common hand disinfection. Similar results were obtained for surgical mask-brand 2 (Figure 4b and Figure 4d). Other methods such as putting hands behind the body can also provide adequate protection of the mask during hand disinfection using alcohol-based sanitizers. The key point is to avoid inhaling alcohol vapor.

**Fig. 4.**
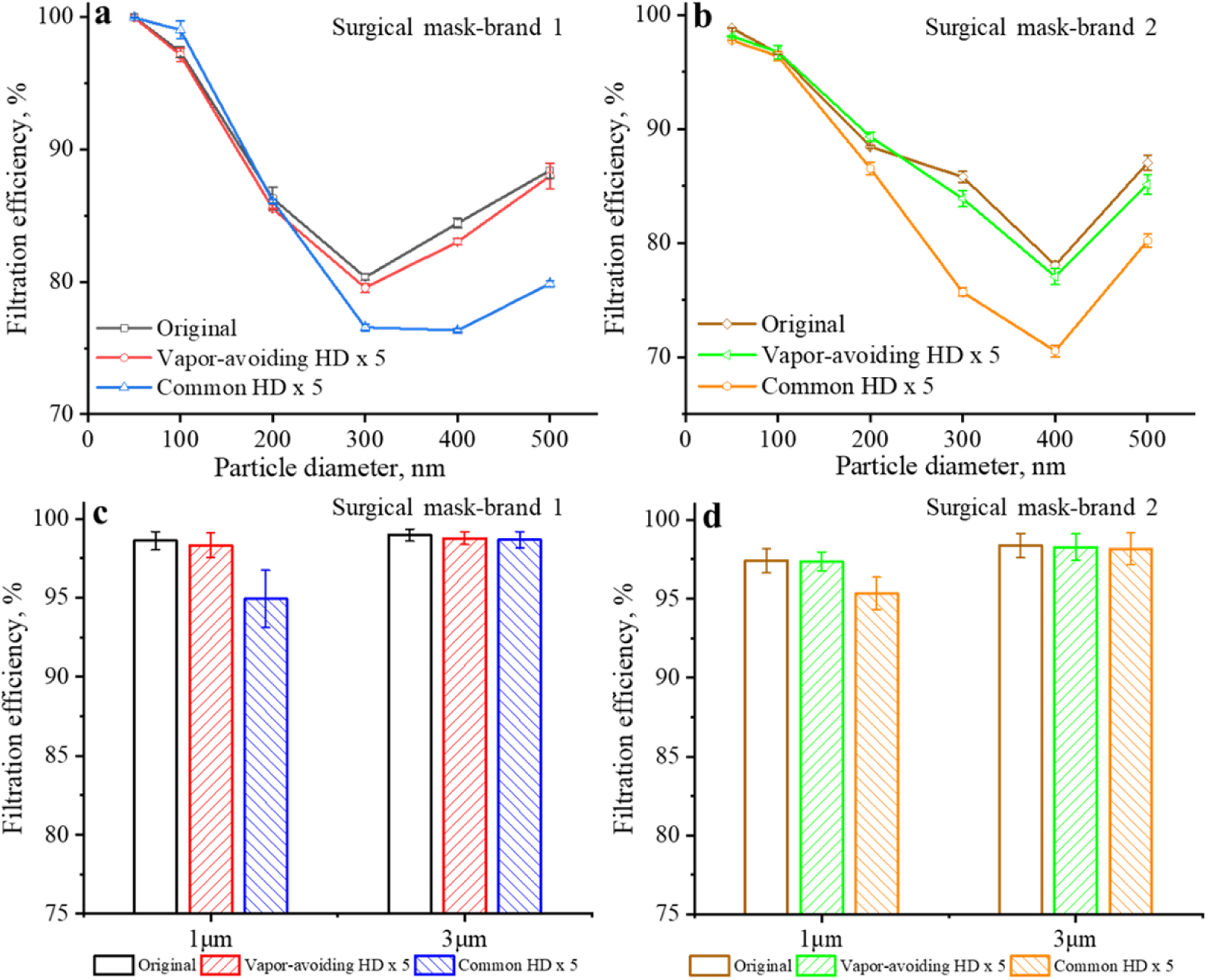
Comparison of the filtration efficiencies of the original surgical masks and surgical masks after 5 times of common hand disinfection and vapor-avoiding hand disinfection; a) and c) Surgical mask-brand 1; b) and d) Surgical mask-brand 2.

In summary, the potential risk of applying alcohol-based sanitizers while wearing masks has been ignored. Using alcohol-based sanitizers for hand disinfection may degrade the filtration efficiencies of the in-use masks, and thereby weaken the protection for the mask wearers when they are exposed to virus-laden aerosols. For a widely used brand of surgical mask, the degradation of the filtration efficiency for 300 nm particles was observed after one time of common hand disinfection. When the number of common hand disinfection increased to five, the filtration efficiencies for 400 and 500 nm particles degraded by more than 8%. In contrast, the alcohol vapor had little influence on the filtration performance of a brand of N95 respirator. Only 0.20±0.01 % of filtration efficiency degradation for 50 nm particles was observed after ten times of common hand disinfection. Compared to the surgical mask, the stronger resistance of the N95 respirator to alcohol vapor was attributed to its thicker outermost layer and multiple charged inner layers. N95 respirators are mainly used by medical staffs in the current situation and surgical masks are widely employed in healthcare settings. High dose of alcohol-based sanitizers is used for not only hand disinfection but also medical device disinfection in hospitals. By applying the precautionary principle in the case of highly dangerous viruses, the influence of alcohol vapor generated during disinfection processes on N95 respirators and surgical masks should be considered, especially in healthcare settings. Cotton masks are not electrostatic filters and their filtration performance is not influenced by alcohol vapor. However, cotton masks may have low efficiencies and are not commonly used by medical personnel.

The common hand disinfection used in the present study followed the standard hand disinfection steps recommended by WHO, and the position of the hands was intentionally kept consistent. The individual differences in the hand disinfection steps and body position may cause different effects on the mask performance than those shown here.

There are no shortcuts and only a comprehensive approach can slow down the spread of the current pandemic. Wearing masks is a critical part of the comprehensive prevention measures, therefore more attention should be paid to the reliability of masks. Using alcohol-based sanitizers for hand disinfection may degrade the filtration performance of masks by dissipating the charges. Vapor-avoiding hand disinfection is a simple and efficient practice to mitigate such risks. We recommend adding the vapor-avoiding hand disinfection in the guide of hand hygiene.

## Supporting information

The details of particle filtration and electrostatic potential tests, and additional figures.

## Data Availability

All data used during the study appear in the submitted article.

## ASSOCIATED CONTENT

### Supporting Information

The details of particle filtration and electrostatic potential tests, and additional figures.

## AUTHOR INFORMATION

### Author Contributions

W.H., Y.G., and J.W. conceived the research ideas and wrote the manuscript. W.H. and Y.G. conducted the experiments. J.L. and Y.Y. contributed to the data analysis. All authors have discussed the results and have given approval to the final version of the manuscripts.

### Notes

The authors declare no competing financial interest.

## ACKNOWLEDGMENT

The work was partially supported by Innosuisse project 46668.1 IP-ENG “ReMask: Strategies for innovations for Swiss masks needed in pandemic situations” and Center for Filtration Research at University of Minnesota. We thank the support of National Science and Technology Major Project of China (Award ID: 2017YFC0211801; 2016YFC0801704; 2016YFC0203701; 2016YFC0801605; 2019JH2/10100004). The authors also thank the financial aid from the project of China Scholarship Council, China.

