## Supplementary material for "Filtration Performance Degradation of In-Use Masks by Vapors from Alcohol-Based Hand Sanitizers and the Mitigation Solutions": The details of particle filtration and electrostatic potential tests, and additional figures.

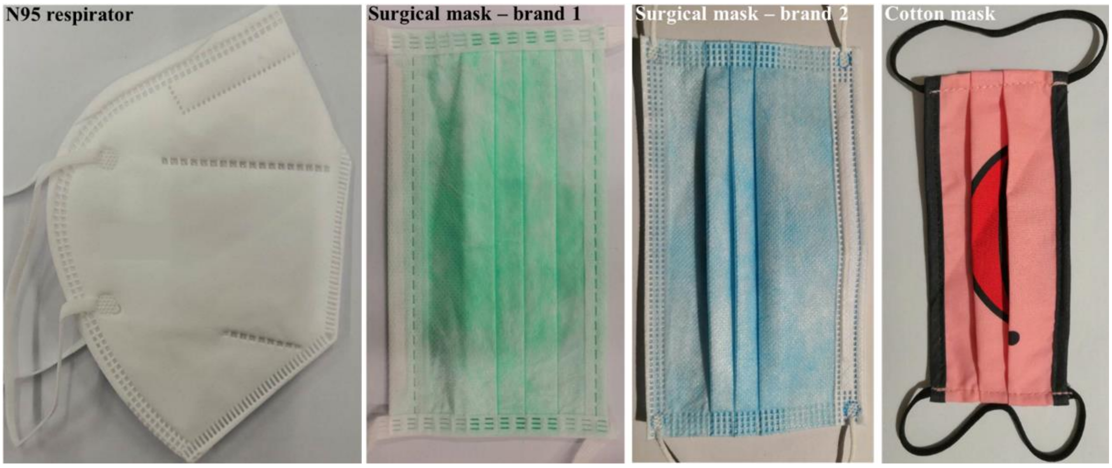


Fig. S1 The N95 respirator, surgical masks (brand 1 and brand 2), and cotton mask used in the present study.

**Particle Filtration Test.** The cut-out samples were circular in shape with a diameter of 4.5 cm, and placed in the filtration setup to measure the efficiency. As shown in Fig. S2a, polydisperse NaCl aerosols were generated by an atomizer (TSI 3079A) and dried by a diffusion dryer. Particles with mobility diameters of 50, 100, 200, 300, 400, and 500 nm were selected using a differential mobility analyzer (DMA, TSI 3081L), and then neutralized using a Kr-85 source. The particle concentrations up-stream and down-stream, which were used to calculate the filtration efficiencies of the test samples, were measured by two condensation particle counters (CPC TSI 3775). A sheath to aerosol flow ratio (SAFR) of 10 in the DMA was applied in order to minimize the artifact due to the multiply-charged particles. For the filtration efficiency test of 1 and 3 μm particles, the monodisperse polystyrene latex (PSL) particles were employed. An aerodynamic particle sizer (APS, TSI 3321) was used to measure the particle concentrations up-stream and down-stream (Fig. S2b). A face velocity of 5.3 cm/s was applied to the mask samples during the filtration test, because it corresponded to a moderate breathing rate (55 L/min) and was a commonly used test velocity for fabric filters and personal protection devices.

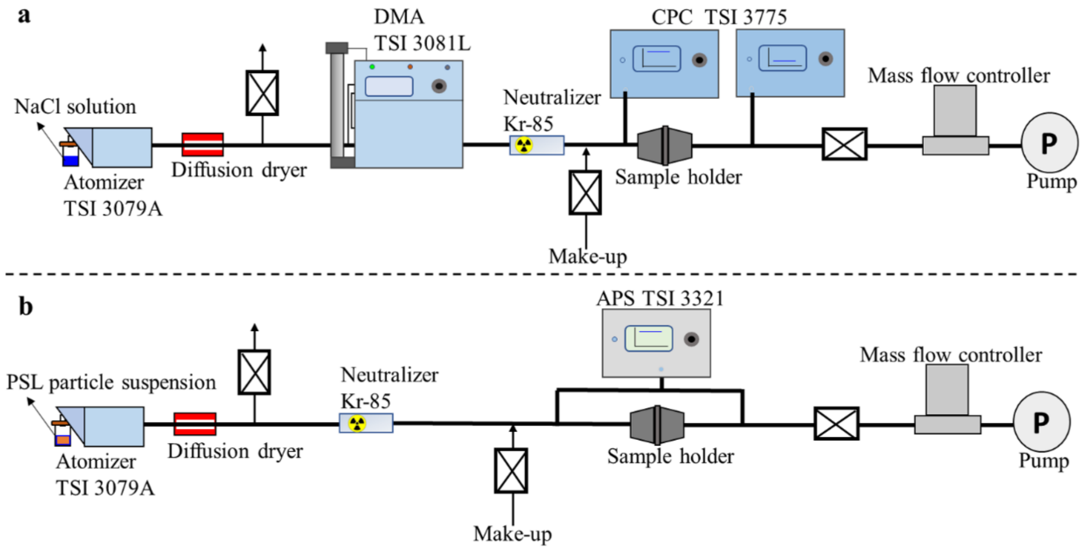


Fig. S2 a) Setup for the NaCl particle filtration test in the size range of 50-500 nm; b) Setup for the PSL particle filtration test in the size range of 1-3 μm.

**Electrostatic Potential Test.** The circular mask samples were put on a grounded metal platform, and the electrostatic potential was measured using an electrostatic voltmeter (Fig. S3). The probe of the electrostatic voltmeter was set at approximately 5 mm above the test sample. The electrostatic potential of the five positions on the sample were measured, and the measurement was repeated for three different pieces.

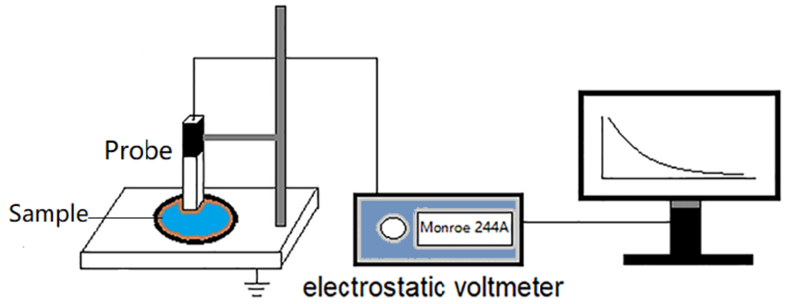


Fig. S3. Schematic of the electrostatic potential test apparatus

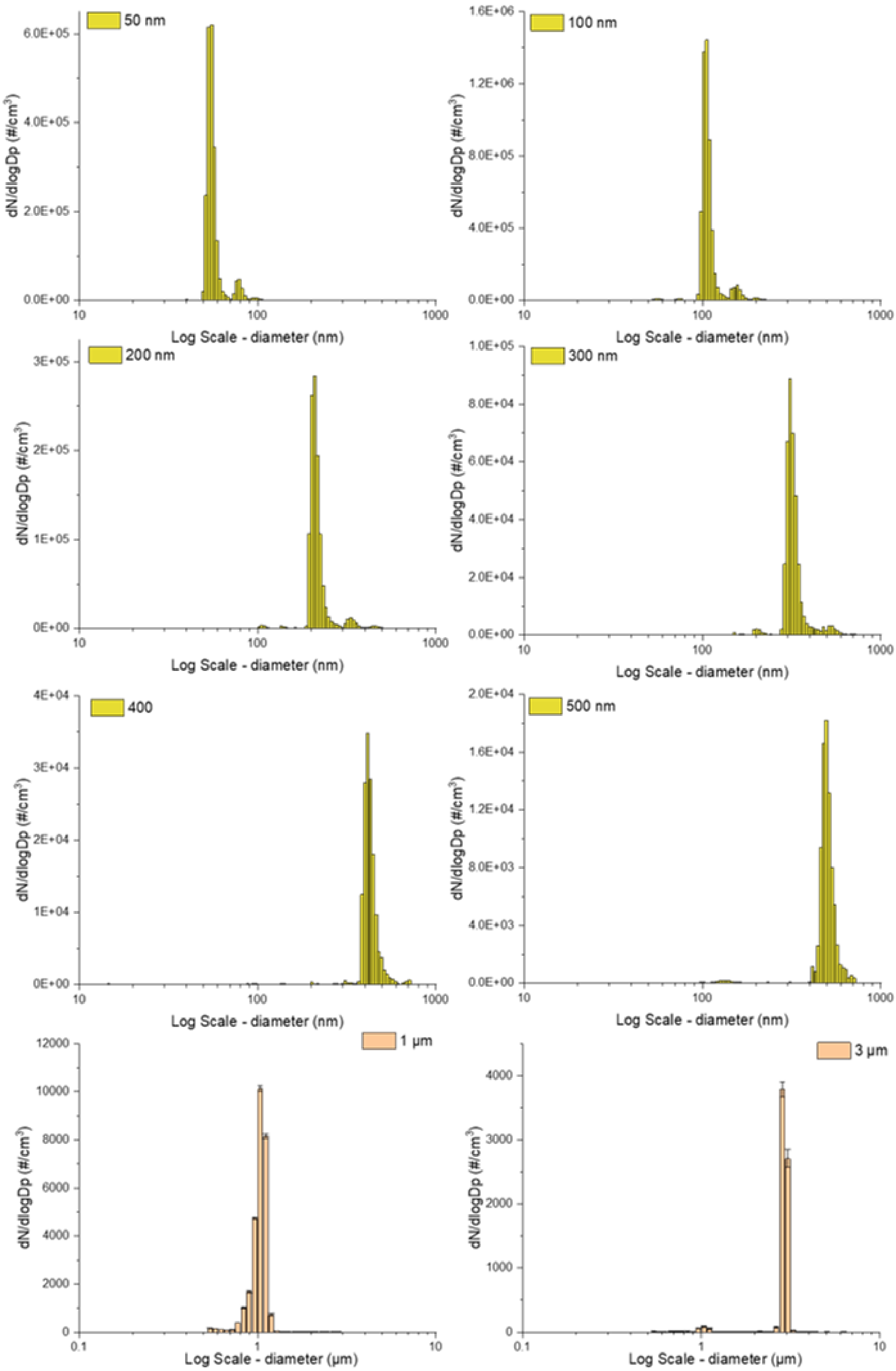


Fig. S4 Size distributions of classified mono-disperse NaCl particles (50-500 nm) and PSL particles (1 and 3 μm) used for the particle filtration test.

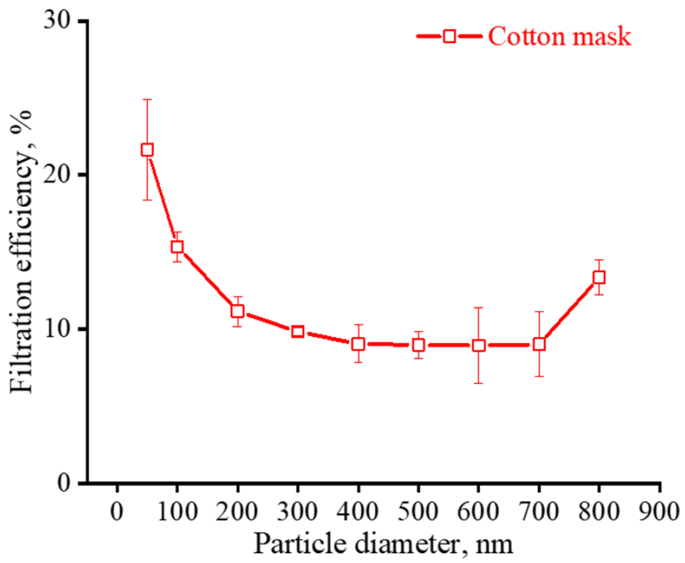


Fig. S5 Filtration efficiencies of the cotton mask for the particles in the range of 50-800 nm.

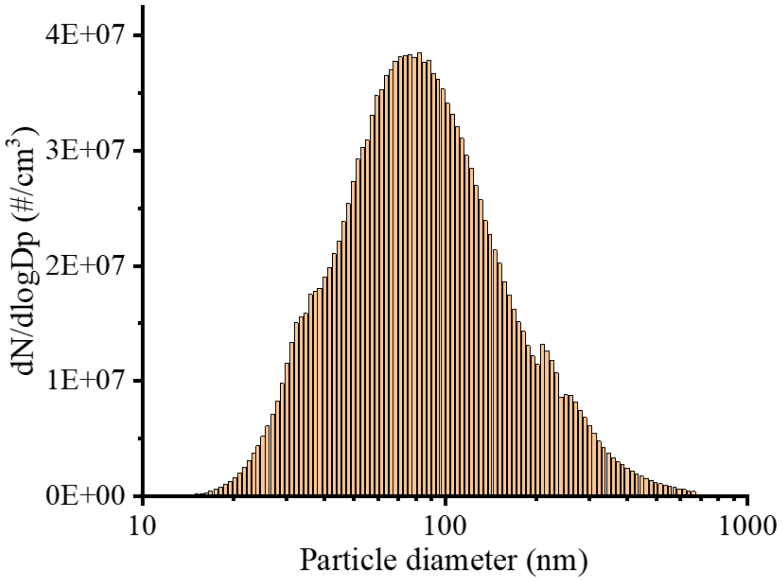


Fig. S6 Size distribution of NaCl particles used for testing the total filtration efficiency of the cotton mask.

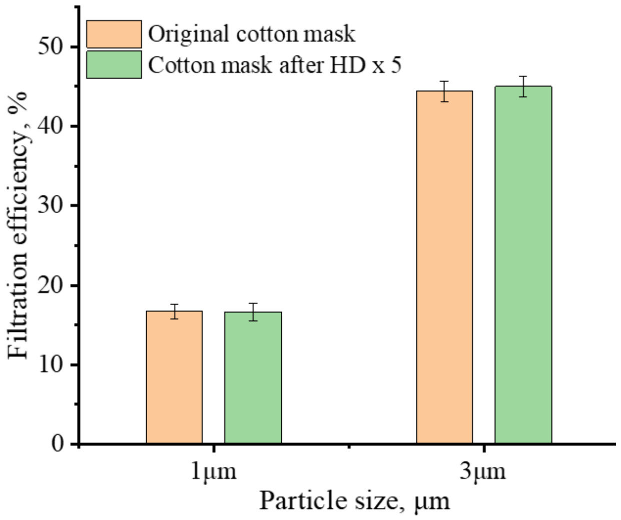


Fig. S7 Filtration efficiencies of brand new and used cotton masks for 1 and 3 μm particles without and with several times of common hand disinfection.

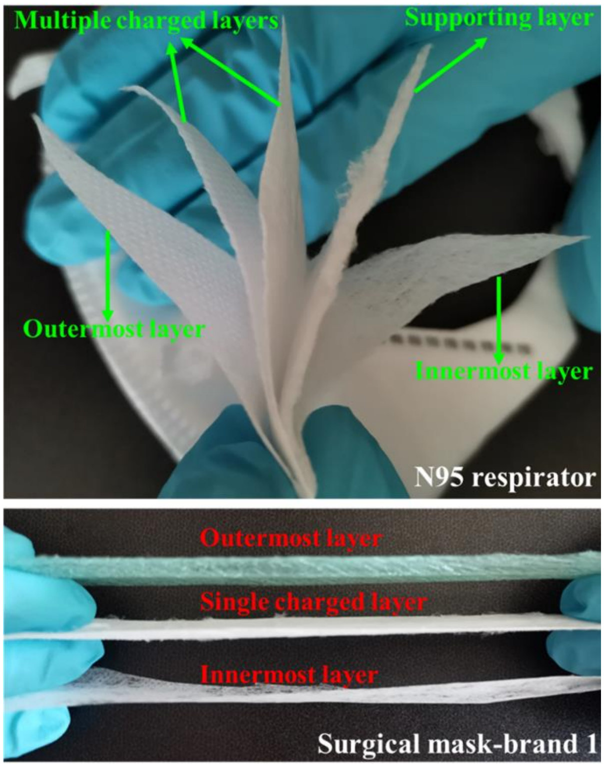


Fig. S8 The structure of multiple layers of the surgical mask and N95 respirator used in the present study.

**Statistical Analysis.** The data of surgical masks were analyzed using Origin 2018 software. Student's *t-test* was used for the electrostatic potential results analysis. A p-value of less than 0.05 indicated a statistically significant difference compared with original surgical mask at a confidence level of 95%. The results are shown in Table S1.

Table. S1 The average electrostatic potential (AVG) of surgical masks after different times of common hand disinfection and the statistical analysis in comparison with the original mask.

| | Original | No HD | HD x 1 | HD x 2 | HD x 3 | HD x 4 | HD x 5 |
|---|---|---|---|---|---|---|---|
| AVG (V) | 744 | 707 | 710 | 702 | 669 | 610 | 589 |
| Uncertainty | 174 | 184 | 182 | 175 | 163 | 154 | 149 |
| *P-value* | Na | 0.28 | 0.29 | 0.25 | 0.11 | 0.02 | 0.007 |
